# Testing federated analytics across secure data environments using differing statistical approaches on cross-disciplinary data

**DOI:** 10.1101/2024.01.06.23300659

**Authors:** S. Gallier, A. Topham, J. Hodson, D. McNulty, T. Giles, S. Cox, J. Chaganty, L. Cooper, S. Perks, P. Quinlan, E. Sapey

## Abstract

**BACKGROUND:** Introducing data-driven technologies into health systems can enhance population health and streamline care delivery. The use of diverse and geographically varied data is key for tackling health and societal challenges, despite associated technical, ethical, and governance complexities. This study explored the efficacy of federated analytics using general linear models (GLMs) and machine learning (ML) models, comparing outcomes with non-federated data analysis.

**METHODS:** A Conditional Transformation Generative Adversarial Network was used to create two synthetic datasets (training set: N=10,000; test set: N=1,000), using real-world data from 381 asthma patients. To simulate a federated environment, the resulting data were distributed across nodes in a Microsoft Azure Trusted Research Environment (TRE). GLMs (one-way ANOVA) and ML models (gradient boosted decision trees) where then produced, using both federated and non-federated approaches. The consistency of predictions produced by the ML models were then compared between approaches, with predictive accuracy of the models quantified by the area under the receiver operating characteristic curve (AUROC).

**FINDINGS:** GLMs produced from federated data distributed between two TREs were identical to those produced using a non-federated approach. However, ML models produced by federated and non-federated approaches, and using different data distributions between TREs, were non-identical. Despite this, when applied to the test set, the classifications made by the federated models were consistent with the non-federated model in 84.7-90.4% of cases, which was similar to the consistency of repeated non-federated models (90.9-91.5%). Consequently, overall predictive accuracies for federated and non-federated models were similar (AUROC: 0.663-0.669).

**INTERPRETATION:** This study confirmed the robustness of GLMs utilising ANOVA within a federated framework, yielding consistent outcomes. Moreover, federated ML models demonstrated a high degree of classification agreement, with comparable accuracy to traditional non-federated models. These results highlight the viability of federated approaches for reliable and accurate data analysis in sensitive domains.

## Introduction

Embedding new, data driven technologies into health and care systems can improve population health and bring efficiencies to health and social care delivery. Correspondingly, digital health innovation is a stated priority for international organisations and governments, such as the World Health Organisation^1^ and the UK Government^2^.

Tackling complex health and societal challenges using data-driven approaches requires access to data from different sectors and geographical sources. However, data access can be challenging. Aside from the technical challenges of integrating large, disparate data, there are ethical and governance restrictions associated with pooling highly sensitive, individual-level data. Recent papers and reviews have recognised the barriers to data egress and have suggested different solutions for analysis, including the concept of federated analytics^3–6^. Federated analytics is a data paradigm that enables different Data Controllers, and those they authorise, to collaboratively perform analytics on their respective local data, without exchanging the raw data itself^7, 8^. In essence, Data Controllers enable the deployment of code across their data with only aggregated results extracted from their local environments in lieu of the data itself.

There are advantages and disadvantages to a federated approach to data analysis. Limited direct exposure to data and a lack of data egress can address some concerns of privacy, security and governance, although model updates and partial aggregates can inadvertently, under certain circumstances, lead to the sharing of personal information^9, 10^. Without direct exposure to the data, analysts are more reliant on the Data Controller or agreed processor to perform data cleansing, and opportunities to explore the data are limited^11^. Also, there are concerns that federated analytics may reduce the accuracy of results in comparison to performing analysis on pooled data^7^.

This study tested the ability to federate analytics utilising differing statistical approaches, namely general linear models (GLMs) and machine learning (ML) models. The resulting outputs were then compared to those produced using non-federated data.

## Methods

This research was conducted with Health Research Authority and Research Ethical Approvals (East Midlands – Derby Research ethics committee, reference 20/EM/0158).

### Sample data

This study used a real-world dataset, from which a synthetic dataset was generated for the purpose of testing federated versus pooled data analysis. The real-world data was from a cohort of 381

patients with a physician-confirmed diagnosis of asthma who were admitted to secondary care at a single centre in 2019. The response variable was readmission within 30 days, which occurred in 12·9% of cases. Clinical data was linked to meteorological and air quality data from The Centre for Environmental Data Analysis (CEDA), for the day of admission using the subjects’ home address for geolocation; see Table 1 for details of the variables included in the real-world dataset. From this two synthetic datasets were generated using a Conditional Transformation Generative Adversarial Network (CT-GAN) deep learning method^12^ from the Synthetic Data Vault (SDV) library, an open-source Python library^13^. The first synthetic dataset was a “training set” of N=10,000 cases, and the second was a “test set” of N=1,000 cases.

**Table 1.** Characteristics of synthetic data used in modelling. Legend. Variable characteristics of the synthetic data used in the modelling provided in category groups. IMD: Index of Multiple Deprivation, O2: Oxygen, PM10: Particular matter 10 particle count, PM2.5: Particular Matter 2.5 particle count.

| Variable | Levels |
| --- | --- |
| **Outcome** |  |
| Readmission within 30 Days | No, Yes |
| **Patient Demographics** |  |
| Age at Index Admission | *Continuous* |
| Sex | Female, Male |
| Ethnicity | White, South Asian, East Asian, Black, Mixed, Other |
| IMD Decile | 1, 2, 3, 4, 5, 6, 7, 8, 9, 10 |
| Height | *Continuous* |
| Weight | *Continuous* |
| **Patient Physiology** |  |
| Asthma Type | SNOMED195967001, J450, J458, J459, J46X |
| Eosinophil Count | *Continuous* |
| Respiratory Rate | *Continuous* |
| O2 Saturation | *Continuous* |
| Peak Flow | *Continuous* |
| **Medications** |  |
| Oral Prednisolone | No, Yes |
| Inhaled Steroids | Beclometasone, Budesonide, Ciclesonide, Duoresp, Fluticasone, Flutiform, Fostair, None, Oxis, Relvar Ellipta, Sereflo, Seretide, Sirdupla, Symbicort, Trelegy |
| **Date of Index Admission** |  |
| Month | 1, 2, 3, 4, 5, 6, 7, 8, 9, 10, 11, 12 |
| Weekday | Working Day, Weekend, Public Holiday |
| Hour of Day | 0, 1, 2, 3, 4, 5, 6, 7, 8, 9, 10, 11, 12, 13, 14, 15, 16, 17, 18, 19, 20, 21, 22, 23 |
| **Atmospheric Conditions at Index Admission** |  |
| PM10 Concentration | *Continuous* |
| PM2.5 Concentration | *Continuous* |
| Nitrous Oxide Concentration | *Continuous* |
| Nitrogen Dioxide Concentration | *Continuous* |
| Sulphur Dioxide Concentration | *Continuous* |
| Temperature | *Continuous* |
| Relative Humidity | *Continuous* |
| Dew Point | *Continuous* |

To simulate a scenario where federated analytics would be applicable, datasets were then divided across “nodes”, each of which was a Microsoft Azure Trusted Research Environment (TRE), and represented the data held in a different database by a different Data Controller (e.g., in different hospitals). In addition, the full dataset was also held, to allow non-federated (“pooled”) analysis to be performed, as the gold standard for comparison.

### Federated learning for general linear models

The algorithms underlying the calculation of the standard deviation, analysis of variance (ANOVA), and Fisher-Neyman factorisation were evaluated, to understand their potential for federation (see Supplementary Analysis 1 for further details). A simple one-way ANOVA was selected to test the federation of a GLM, with the resulting model being compared to one produced using a non-federated approach. This used the test set of N=1,000 cases, with the 10-micron particulate matter (PM10) counts as the dependent variable, and readmission within 30 days as a (binary) independent variable. For the models on federated data, the dataset was divided between two simulated nodes using four different approaches to simulate different scenarios; see Figure 1. Specifically, the “row-split” randomly divided the cases in a 1:1 ratio between the two nodes, to simulate the situation where each node held data for a different set of cases, but the same set of variables. The “column-split” divided the data such that the dependent and independent variables were on different nodes, to simulate the situation where each TRE held data for all cases, but for a different set of variables. These two approaches were also combined into a “row-and column-split” and “ragged-split”, where each TRE held data for different sets of cases, and different sets of variables for these.

**Figure 1.**
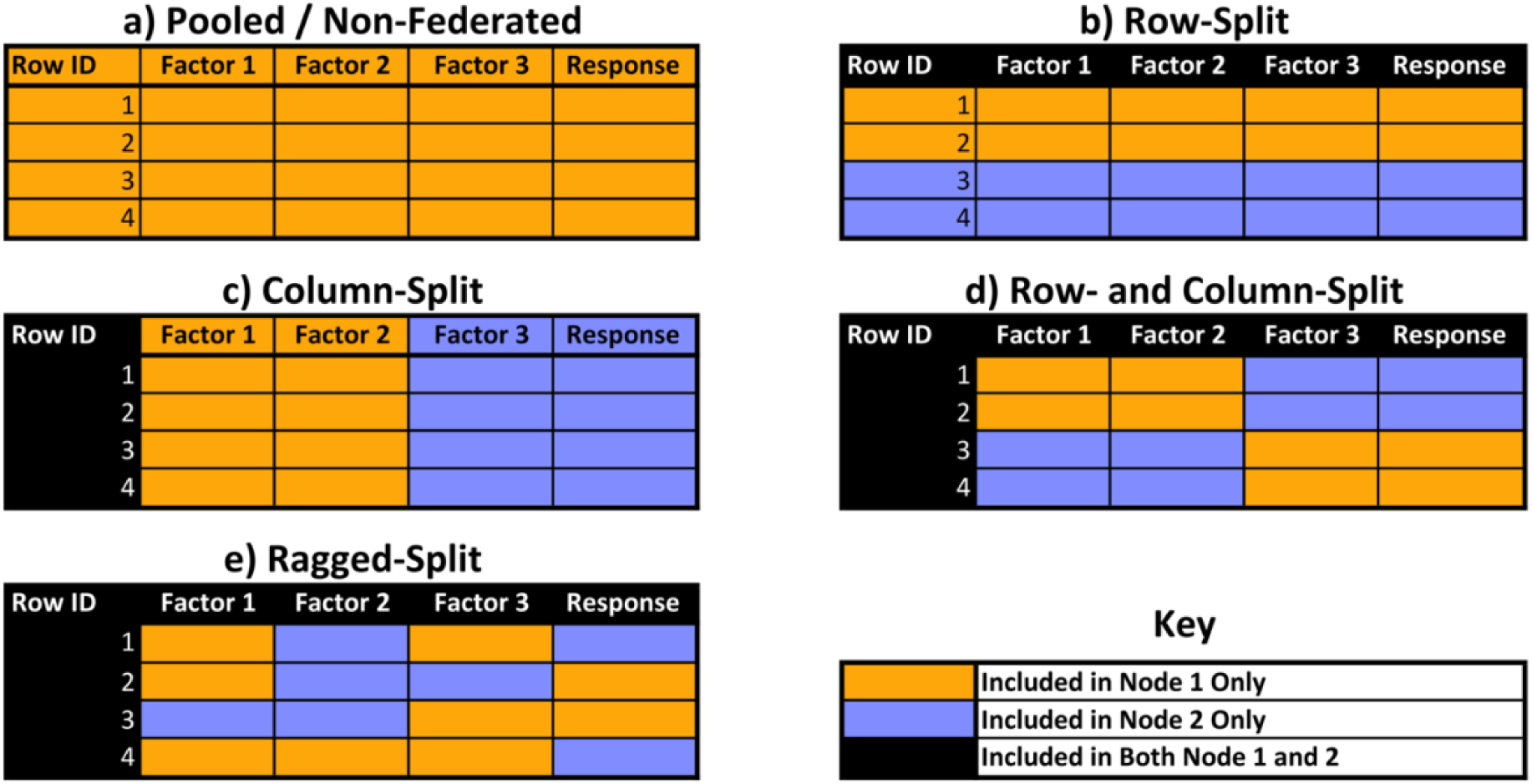
Approaches to splitting data across federated models. Legend. Each panel represents a different approach to dividing the rows (cases) and columns (variables) of a table across two nodes, with the cell colours indicating the node on which data for a specific cell would be held. Figure a) represents pooled/non-federated data, where the whole table is held on a single node; b) represents a row-split, where cases are divided between the two nodes, both of which include all variables; c) represents a column-split, where variables are divided between the two nodes, both of which include all cases; d) represents a combination of a row- and column-split, where subsets of variables and cases are held on different nodes; and e) represents a ragged-split, where individual variables*case combinations can be held on either node.

For the federated models, in addition to the TRE nodes, there was also a controlling hub, which sent requests for data to the individual TRE nodes and compiled the resulting data to perform the ANOVA analysis. The controlling hub never directly accessed the underlying datasets, with TRE nodes instead only sharing aggregated data, including sums, and row counts, which could be used to compute global means and degrees of freedom accurately (see Figure 2).

**Figure 2.**
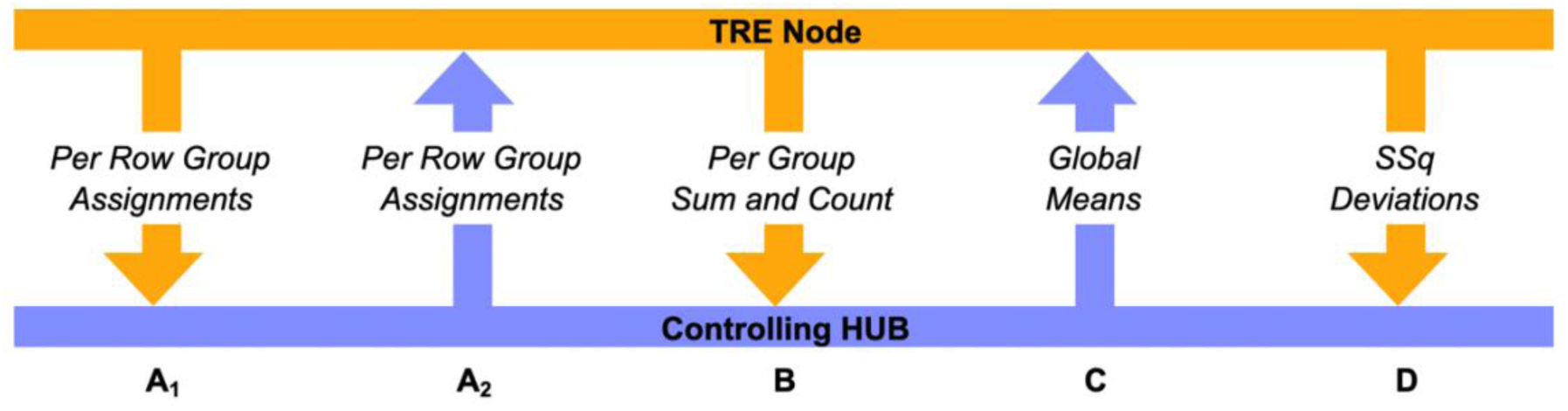
Data communications in a federated ANOVA environment. Legend. Hub-to-node communications are indicated by blue arrows, and node-to-hub communications by orange arrows. A_1_ and A_2_ are necessary for column-split, a combined row- and column-split, and ragged data partitioning, where each node has incomplete sight of the covariates for each row, and so it is necessary for the hub to retrieve the known covariates from each node, combine them and distribute the complete set back to each node, thus relying on covariates not being sensitive in nature. In all partitioning modes, for every group of covariates, the nodes can return the sum of the response and row count (B), as well as being able to compute and return the sum squared deviation (D) from a given global mean computed by the hub using the per-group sum and row counts (C). API: Application Programming Interface, TRE: Trusted Research Environment, SSq Deviations: Sum of Squared Deviations.

In scenarios where the data were partitioned among TRE nodes in a column-wise manner, either solely or in combination with row-wise partitions, unique identifiers were established for each row. The TRE nodes then shared the factors assigned to each unique row throughout the network (Figure 2, steps A_1_ and A_2_) to produce the statistical outputs.

Once the unique row identifiers and factor assignments were established, the following steps were shared across all partitioning modes:

1. Calculation of Sum and Row Counts: Each TRE node returns the sum and row counts for each combination of factors to the controlling hub. This step allows for the computation of group-wise and global means of the response variable, as well as various degrees of freedom across the federated data set (Figure 2, step B).
2. Distribution of Global Mean: The global mean is sent back to each TRE node (Figure 2, step C).
3. Computation of Sum of Squared Deviations: Each TRE node calculates the sum of squared deviations from the global mean and returns it to the controlling hub (Figure 2, step D).
4. Calculation of ANOVA Statistics: The final ANOVA statistics, along with confidence intervals for the differences between means, are computed using the sum of squared deviations and Tukey’s HSD respectively.

### Federated learning for machine learning models

The performance of ML models using existing software stacks, trained using a federated approach was assessed. For this analysis, only the row-split scenario was considered, with the cases from the training set (N=10,000) being randomly divided between three TRE nodes in a 5:3:2 ratio. We tested two different modes of federation, which are visualised in Figure 3. The first was “online” learning, where a single ML model is initialised and passed sequentially between all federated environments. The second approach was “concurrent” learning, where separate models are trained in each federated environment and returned to the controlling hub, where they are combined or averaged. The response variable was 30-day readmission, with all 24 factors in Table 1 considered as predictors. ML models comprised Gradient Boosted decision trees from CatBoost 1.2 (Yandex, Moscow, Russia), a high-performance open-source library^14, 15^. An Application Programming Interface (API) endpoint was utilised to execute a training cycle using the concurrent mode of federation. All models were generated using the same random seed, in order to negate the impact of this when comparing across models. However, to test the potential impact that changing the random seed could have, the pooled (i.e., non-federated) model was also repeated a further three times using different random seeds, which were compared to the original pooled model.

**Figure 3.**
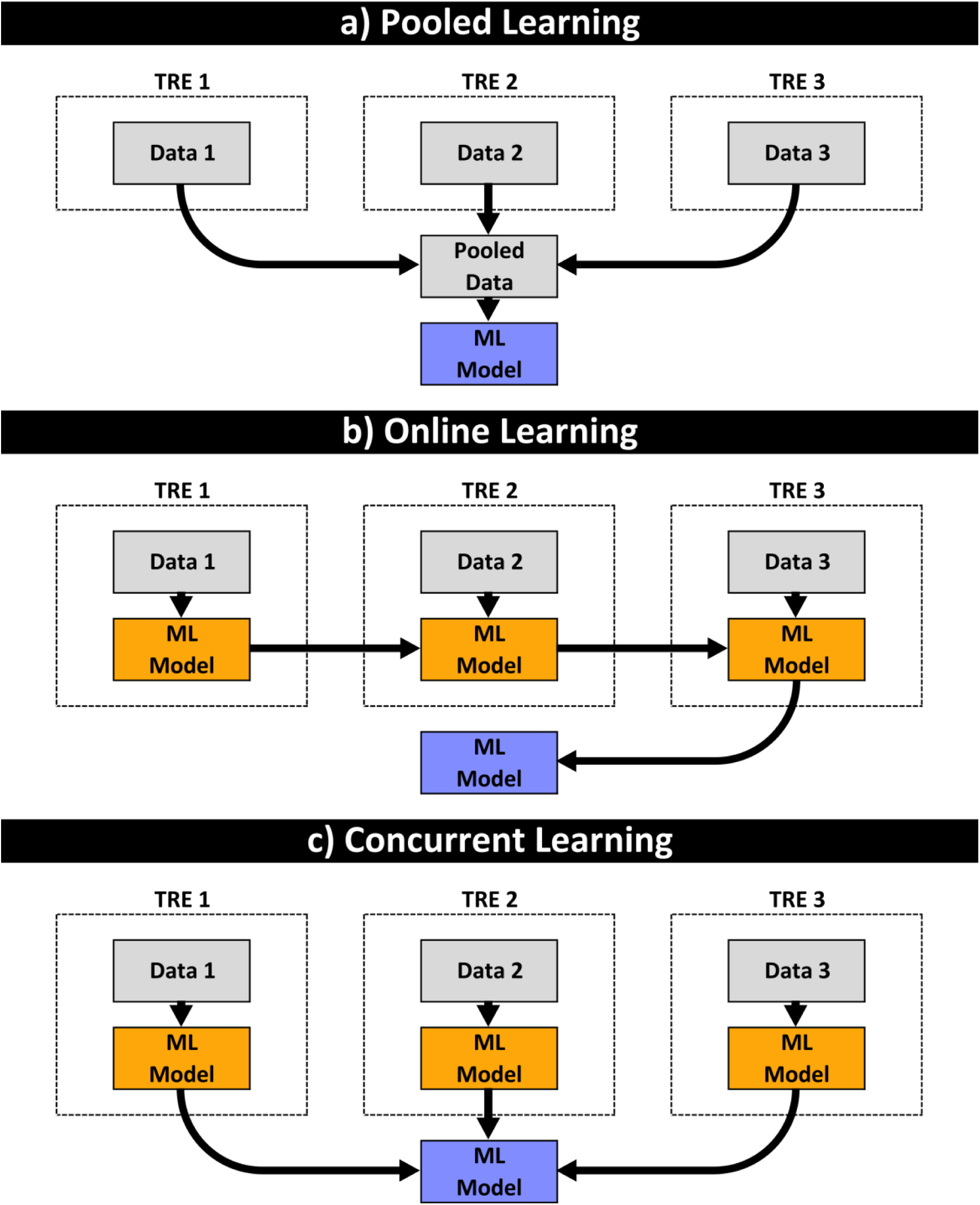
Schematic of the learning methods utilised for federated machine learning. Figure a) illustrates pooled (i.e., non-federated) learning, where data are moved from each TRE node into a central pooled environment for analysis by the ML algorithm. For the federated approaches, the federated learning system first passes an ML model into each TRE node. In online learning (Figure b), this ML model is trained iteratively across each TRE node, with each node performing local model updates using its own data, before sending only the updated model parameters to the next node, with the final node sending the final model parameters to a central server. In concurrent learning (Figure c), the ML model training is performed in parallel across all nodes, before updating the global model simultaneously. ML: Machine Learning, TRE: Trusted Research Environment.

The resulting models were then applied to the test set (N=1,000), to compare consistency and performance. Each model was used to produce both a predicted probability of 30-day readmission, and a predicted binary classification for this outcome for each case. The predicted probabilities were compared between models using Spearman’s rho, and the binary classification using the percentage agreement, in order to test for consistency of the models. In addition, the predictive accuracy of each model was quantified using the area under the receiver operating characteristic curve (AUROC) for the predicted probability, and the accuracy, sensitivity, specificity and positive/negative predictive value statistics for the binary classifications. The AUROCs were then compared between the models using the algorithm suggested by Delong et al., with p<0.05 deemed to be indicative of a statistically significant difference in performance^16^.

## FINDINGS

### Federation of general linear models

ANOVA models were produced for the pooled data, as well as on data federated across two TREs, using the four approaches detailed in Figure 1. All five of these models returned identical results, which are reported in Table 2.

**Table 2.**
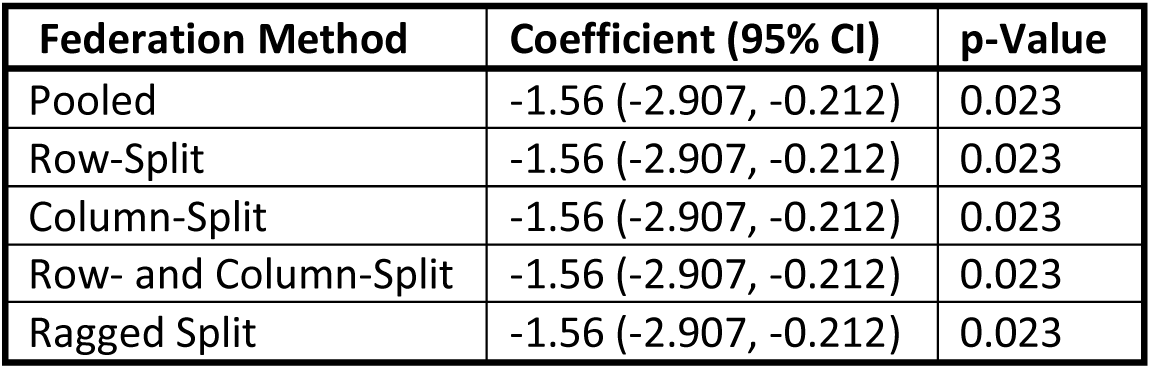
Comparison of ANOVA models across pooled and federated methods. Legend. Results are from ANOVA modelling on the synthetic dataset, with PM10 concentration as the dependent variable, and readmission within 30 days as a (binary) independent variable. The coefficient represents the difference between the 30 day readmission yes versus no groups, and is reported alongside the 95% confidence interval (95% CI) computed using Tukey’s HSD. Separate models were produced using non-federated data (“pooled”), as well as using the four approaches to federation described in Figure 1. PM10: Particular matter 10 particle count, HSD: honesty significance difference, CI: Confidence Interval.

### Federation of machine learning models

ML models were then trained on the training dataset, both using a pooled approach, and with data federated across three TREs using a row-split approach; the federated models were produced using both concurrent and online learning approaches (as per Figure 3). Applying the resulting models to the test set for validation found that the classifications made by pairs of models to be in agreement in between 84·7-90·4% of cases, with Spearman’s rho for the predicted probabilities ranging from 0·844-0·938 (Table 3). The pooled (i.e., non-federated) model was also repeated a further three times, with each using a different random seed, in order to assess how this would impact the resulting models. The agreement between the classifications produced by these models and the primary pooled model ranged from 90·9-91·5%, with Spearman’s rho for the predicted probabilities ranging from 0·914-0·931. As such, the consistency of classifications made between ML models trained on differently federated data was only marginally lower than that stemming from training repeated ML models on identical, non-federated datasets.

**Table 3.** Comparison of classifications made by machine learning models. Legend. Results are from machine learning models trained on a non-federated (“pooled”) or federated (“concurrent” or “online”) synthetic dataset, for the prediction of 30-day readmission. The resulting models were then applied to the test set (N=1,000), and the binary classifications made by each pair of models were compared. *The Spearman’s rho correlation coefficient between the predicted probabilities produced by the two models.

| Pooled versus Online<br>(Agreement: 84.7%; rho: 0.844*) |  |  |  |
| --- | --- | --- | --- |
|  |  | Online |  |
|  |  | Classification: No | Classification: Yes |
| Pooled | Classification: No | 573 | 99 |
|  | Classification: Yes | 54 | 274 |
| Pooled versus Concurrent<br>(Agreement: 90.4%; rho: 0.904*) |  |  |  |
|  |  | Concurrent |  |
|  |  | Classification: No | Classification: Yes |
| Pooled | Classification: No | 652 | 20 |
|  | Classification: Yes | 76 | 252 |
| Online versus Concurrent<br>(Agreement: 87.7%; rho: 0.938*) |  |  |  |
|  |  | Concurrent |  |
|  |  | Classification: No | Classification: Yes |
| Online | Classification: No | 616 | 11 |
|  | Classification: Yes | 112 | 261 |

Despite the differences between the binary classifications made by the federated and non-federated approaches, the pooled, online and concurrent models had similar predictive accuracies when applied to the test set (Table 4). The overall accuracy statistic ranged from 64·4%-71·7% and the AUROC for the predicted probabilities ranged from 0·663-0·669. Specifically, the AUROC for the pooled model was 0·667 (95% CI: 0·621-0·713), which did not differ significantly from the 0·663 (95% CI: 0·616-0·710) for the online model (p=0·739) or the 0·669 (95% CI: 0·623-0·716) for the concurrent model (p=0·824).

**Table 4.**
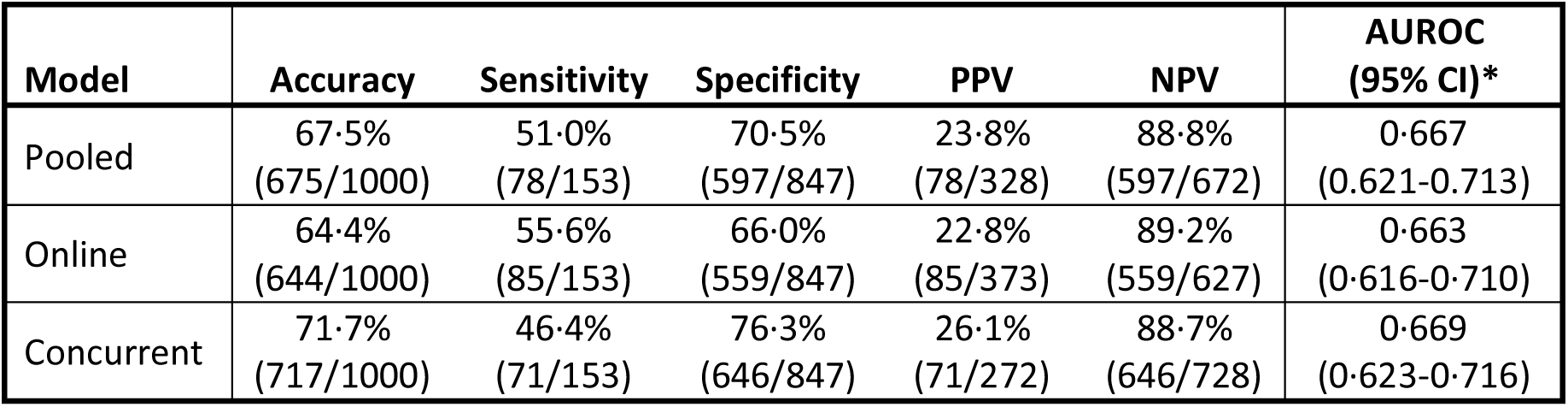
Comparison of predictive accuracy of machine learning models. Legend. Results are from machine learning models trained on a non-federated (“pooled”) or federated (“concurrent” or “online”) synthetic dataset, for the prediction of 30-day readmission. The resulting models were then applied to the test set (N=1,000), and the accuracy of the predictions made by each model was assessed. *The area under the receiver operating characteristic curve (AUROC) was calculated using the predicted probabilities from each model. P(N)PV: Positive (Negative) Predictive Value.

## INTERPRETATION

Federated analytics and learning is increasingly prevalent and may be default methodology for analysing sensitive health data^17–19^. However, the security benefits this offers^20^ can only be realised if the resulting analysis accurately reflects the pooled data analysis.

This study demonstrates the feasibility of employing various statistical approaches within a federated analytics framework across diverse data domains. Selected traditional statistical methods, namely ANOVA, produced identical outputs irrespective of the way the data were partitioned between TRE nodes, which were in turn identical to the non-federated analysis of the pooled dataset. Therefore, for studies requiring analysis using GLM models, federated analytics can be used without impacting the results of the analysis^21, 22^.

The federated analytics for ML models found differences between the models resulting from federated and non-federated approaches, and between the different learning methods used in the federated approaches. The classifications made by the three approaches were generally consistent, with the level of agreement between models being only marginally lower than for repeated models trained on identical non-federated datasets. In addition, the predictive accuracies of the ML models produced using pooled, online, and concurrent models were similar, ranging from 64·4% to 71·7%.

However, it is important to recognise that differences exist in the ML models generated using a federated versus pooled approach. The clinical relevance of the differences in ML models generated using a federated versus pooled dataset, and the resulting impact on the performance of a tool at the individual level will be context dependent. This may be of relevance to regulators, especially for tools used as a medical device, and should be considered in regulatory pathways. Although several aggregation strategies for federated learning knowledge have been proposed, the field is still in its early stages of development, and more work is needed to determine the impact on the models derived. As well as the potential differences in ML models generated using a federated approach, there are other challenges with federated analytics which need to be explored.

As well as the potential differences in ML models generated using a federated approach, there are other challenges with federated analytics which need to be explored. Primarily, it is vital to ensure that data are consistent and accurately matched across nodes. This results in a heightened reliance on metadata, which must accurately reflect the definitions and coding of variables. In addition, the responsibility of data cleansing and harmonisation resides with data controllers, creating potential inconsistencies in data quality across nodes. The lack of standardised frameworks and common data models can also exacerbate difficulties in conducting analytics across varied datasets. The levels of data consistency, cleansing or harmonisation are difficult to identify where a federated approach is used, since no one individual has access to the whole dataset, potentially propagating misinterpretations and flawed analyses where these are inadequate. Secondly, whilst the federated approach can be effective in addressing privacy concerns, sophisticated strategies are required to balance privacy preservation with effective analysis. The governance associated with federated analytics is still in development^23, 24^.

In conclusion, we demonstrate the potential of federated approaches in maintaining predictive accuracy while preserving data privacy and security. For traditional statistical techniques (here, a one-way ANOVA), the models generated using federated approaches were identical to those generated using a non-federated approach, irrespective of how the data were split across nodes. For ML analyses, there was some variability in the models produced using the federated and non-federated approaches, and across the different federated approaches. Although there was no statistically significant difference in predictive accuracy observed between the models, the impact on clinical performance may be context specific and warrants further exploration.

## Supporting information

Supplementary Analysis 1

## Data Availability

To facilitate knowledge in this area, the synthetic data and a data dictionary defining each field will be available to others through application to PIONEER via the corresponding author.

https://catalogue.ceda.ac.uk/

## Acknowledgements

This work was supported by PIONEER, the Health Data Research Hub in acute care and funded by Innovate UK. This work uses synthetic data, however this was generated based on data provided by patients and collected by the NHS as part of their care and support. We would like to acknowledge the contribution of all staff, key workers, patients and the community who have supported our hospitals and the wider NHS.

## AUTHOR CONTRIBUTION

SG and ES designed the study, collated data, performed some analysis, wrote the manuscript. JH, DMcN and AT curated data and supported statistical analysis. SG performed statistical analysis and wrote the first draft of manuscript. All authors amended the manuscript and approved the final version.

