## Supplementary Analysis 1 for "Testing federated analytics across secure data environments using differing statistical approaches on cross-disciplinary data"

### SUPPLEMENTARY ANALYSES 1

### Statistical Results

Two simple examples are the mean $\mu$ and variance $s^{2}$. The mean is simply a set of sums and counts and may be partitioned into $p$ smaller sums.

$$\mu= \frac{1}{N}\sum_{i=1}^{N} x_{i}= \frac{1}{\sum_{j=1}^{p} n_{i}} \sum_{j=1}^{p} \sum_{i=1}^{n_{j}} x_{i,j}$$

Where $N=\sum_{j=1}^{p} n_{j}$ and $n_{j}>0$

Likewise the variance may for illustrative purposes be written

$$s^{2}= \frac{1}{N-1}\left[ \sum_{i=1}^{N} x_{i}^{2}-\frac{1}{N}\left( \sum_{i=1}^{N} x_{i} \right)^{2} \right]$$

Each sum within the formulation may be partitioned in its own right.

More generally the Fisher-Neyman factorisation theorem may be applied to the likelihood of generalised linear models having errors from the exponential family. Given a sample $\left\{ X_{i} | i=1 to n \right\}$ with probability density $f\left( x | \theta\right)$where $\theta i$s a parameter vector, the likelihood of $\theta$ given $\left\{ X_{i} | i=1 to n \right\}$ is written.

$$\prod_{i=1}^{n} l\left( \theta| X_{i} \right)$$

Where $l\left( \theta| X_{i} \right)= f\left( X_{i} | \theta\right)$

For data having an exponential family distribution

$$f\left( x | \theta\right)=exp\left[ c\left( \theta\right).T\left( x \right)+d\left( \theta\right)+S\left( x \right) \right]$$

The log likelihood is

$$c\left( \theta\right)\sum_{i=1}^{n} T\left( X_{i} \right)+n.d\left( \theta\right)+\sum_{i=1}^{n} S\left( X_{i} \right)$$

The term $\sum_{i=1}^{n} T\left( X_{i} \right)$ is known as a sufficient statistic for $\theta$. All the information required to calculate $\theta$ is contained within $\sum_{i=1}^{n} T\left( X_{i} \right)$. As shown with the mean, sums of quantities are readily federated.

There is no need to demonstrate federation for Bayesian statistics as Bayesian updates may be applied sequentially in any order with the posterior at step $n$ becoming the prior at step $n+1$.
